# Impact of Cerebrospinal Fluid Leukocyte Infiltration and Neuroimmmune Mediators on Survival with HIV-Associated Cryptococcal Meningitis

**DOI:** 10.1101/2024.05.29.24308130

**Authors:** Samuel Okurut, David R. Boulware, Yukari C. Manabe, Lillian Tugume, Caleb P. Skipper, Kenneth Ssebambulidde, Joshua Rhein, Abdu K. Musubire, Andrew Akampurira, Elizabeth Okafor, Joseph O. Olobo, Edward N. Janoff, David B. Meya, ASTRO Trial Team

## Abstract

**Introduction:** Cryptococcal meningitis remains a prominent cause of death in persons with advanced HIV disease. CSF leukocyte infiltration predicts survival at 18 weeks; however, how CSF immune response relates to CSF leukocyte infiltration is unknown.

**Methods:** We enrolled 401 adults with HIV-associated cryptococcal meningitis in Uganda who received amphotericin and fluconazole induction therapy. We assessed the association of CSF leukocytes, chemokine, and cytokine responses with 18-week survival.

**Results:** Participants with CSF leukocytes ≥50/µL, had higher probability 68% (52/77) of 18-week survival compared with 52% (151/292) 18-week survival in those with ≤50 cells/µL (Hazard Ratio=1.63, 95% confidence intervals 1.14-2.23; p=0.008). Survival was also associated with higher expression of T helper (Th)-1, Th17 cytokines, and immune regulatory elements. CSF levels of Programmed Death-1 Ligand, CXCL10, and Interleukin (IL)-2 independently predicted survival. In multivariate analysis, CSF leukocytes were inversely associated with CSF fungal burden and positively associated with CSF protein, interferon-gamma (IFN-γ), IL-17A, tumor necrosis factor (TNF)-α, and peripheral blood CD4^+^ and CD8^+^ T cells expression.

**Conclusion:** 18-week survival after diagnosis of cryptococcal meningitis was associated with higher CSF leukocytes at baseline with greater T helper 1 (IFN-γ, IL-2 and TNF-α cytokines), T helper 17 (IL-17A cytokine) and CXCR3^+^ T cell (CXCL10 chemokine) responses. These results highlight the interdependent contribution of soluble and cellular immune responses in predicting survival with HIV-associated cryptococcal meningitis.

## 1. Background

Cryptococcal meningitis (CM) remains one of the leading causes of AIDS-related death worldwide[1–3]. Cryptococcal infection of the central nervous system (CNS) leads to activation of CNS-resident and patrolling immune cells [4–6]. Activated leukocytes in cerebrospinal fluid (CSF) produce chemokines, cytokines, and other immune-mediating factors responsible for shaping infection, CNS inflammation and disease outcome [7,8]. The evoked immune response contributes to cryptococcal killing by anti-fungal medications and influences cryptococcal fungal burden and host survival [9–11]. Low numbers of leukocytes in CSF are a harbinger of immune suppression and cryptococcal-related mortality [12,13]. Monocytes, the precursors of macrophages that harbor intracellular replication machinery for *Cryptococcus*, show impaired immune activation and function among patients with cryptococcal meningitis with lower probability of survival [14,15]. Increased cryptococcal phagocytosis by macrophages is associated with increased intracellular fungal replication, leading to high CSF cryptococcal fungal burden and lower probability of survival [15]. These earlier observations suggest an influence of cryptococcal and/or other underlying immune suppressive factors that selectively evade T helper (Th) -mediated defense. The evasion of the T cell helper function allows unchecked intracellular fungal replication leading to high fungal burden among patients and a lower probability of recovery.

The immune-activated cytokine, chemokine, and checkpoint regulatory responses are important in orchestrating sequestration of *Cryptococci* in the CNS. The Th1 cytokine interferon (IFN)-γ elicits signal transduction to activate intracellular pathogen killing by infected macrophages [16]. The CXCL10-interferon inducible protein 10 (IP-10) supports the recruitment of activated CXCR3^+^ T cells and Natural killer (NK) and NK T cells to mediate a Th1-induced immune response [17]. Women with low CSF levels of the CXCR3^+^ T cell chemoattractant chemokine, CXCL10 and Th17 T cell-activating cytokine, IL-17A were more likely to die on anti-fungal therapy [18]. These observations suggest immune-mediated evasion pressure by such factors that impair cryptococcal host recovery, as seen among patients with high fungal burden and diminished immune response.

Casadevall and Pirofski’s immune-pathogen damage response framework suggests that an optimal treatment strategy for infectious pathogens should enhance pathogen killing and control the potential detrimental effect of host-induced immune response to infection[19]. However, striking a balance between immune- and antifungal-mediated cryptococcal killing while limiting bystander neuroimmunopathology is a challenge, especially among individuals with severe immune deficiency [20–22]. To understand immune-associated survival mechanisms with infiltrating CSF leukocytes, we examined whether the expression of CSF Th1, Th17 cytokines, and chemokine responses correlate with levels of CSF infiltrating leukocytes, CSF fungal burden, and host 18-week survival after diagnosis or study enrolment.

## 2. Methods

### 2.1. Parent trial, participants, site, and setting

We enrolled 401 consenting adults, 241 males and 160 females, with a median age of 35 years, (interquartile range (IQR); 29-40 years) with cryptococcal meningitis from Mulago and Kiruddu Hospitals in Kampala and Mbarara Regional Referral Hospital in Mbarara, Uganda between March 2015 and May 2017. Participants were recruited during the Adjunctive Sertraline for the Treatment of CM trial (ClinicalTrials.gov: NCT 01802385) [23,24]. The CSF cryptococcal antigen (CrAg) lateral flow assay was used to diagnose enrolled patients (Immy Inc., Norman, Oklahoma, USA)[25–27]. Participants or their surrogates provided written informed consent. The Mulago Hospital Research Ethics committee and University of Minnesota institutional review board approved the study.

The CSF was collected by lumbar puncture. CSF leukocytes were counted in fresh CSF using a hemocytometer. CSF was centrifuged at 500g for 5 minutes and the supernatant separated and cryopreserved at -80 ^0^C before thawing for batch testing.

### 2.2. Study design

This was an exploratory study, designed to investigate the association between the levels of CSF leukocyte infiltration demonstrated by the CSF white cell counts and the associated immune responses at CM diagnosis and 18-week survival after diagnosis or enrolment. Participants were systematically selected and stratified by CSF leukocyte number: ≤50/μL, 51-200 cells/μL, and 201-500 cells/μL.

### 2.4. Luminex cytokine and chemokine immunophenotyping

The baseline CSF cytokine and chemokine levels were measured using 1:2 dilutions with a human XL cytokine discovery kit per the manufacturer’s instructions (R&D, Minneapolis, MN). The Luminex CSF data acquisition was performed at the University of Minnesota earlier explained[18,28]. Briefly, the Th1 cytokines regulated through T-bet and STAT1 transcription factors were TNF-α, IFN-γ, IL-2, and IL-12p70. The Th2 cytokines regulated through Gata 3 and STAT 6 transcription factor-modulated cytokines were IL-4 and IL-13. The T follicular helper adaptive cells activating cytokines regulated through Bcl-6 and STAT 3 transcription factors were IL-6 and IL-10. The Th17 cytokines include IL-17A. The innate-like cytokines were IL-15, IL-8/CXCL8 or CXCL8, IL-1RA or IL-1F3 produced by innate lymphoid and myeloid cells among neutrophils, monocytes, macrophages, dendritic cells to mediate cellular chemoattraction to neuroinflammation.

The chemokines that work in synergy with induced cytokines and among activated cells to attract B cells, T cells, and innate lymphoid cells resulting in neuroinflammation were: CXCR3^+^ T cell activating chemokine CXCL10/IP-10 secreted by monocytes, macrophages, dendritic cells, and in the CNS secreted by microglia cells and astrocytes for lymphocyte chemoattraction to neuroinflammation. The CCL11/Eotaxin for myelocyte chemoattraction to neuroinflammation. The IL-8/CXCL8 for neutrophil activation and chemoattraction to neuroinflammation. The immune checkpoint inhibitor was PD-L1/B7-H1 for control of the resultant immune response.

### 2.5. Statistical analysis

Data were analyzed using GraphPad Prism version 9.3.0 (San Diego, California, USA). In the univariate analyses, continuous variables were analyzed using the Mann-Whitney non-parametric unpaired t-test for comparison of sample medians. The difference in survival (binary outcome) was determined using univariate Log-Rank test and multivariate Logistics Regression analysis. The 7.9% (32/401) missingness in survival were systematically imputed using either all alive or dead approach. Kruskal Wallis test (Analysis of variance; ANOVA) was used for simple linear group-wise analysis. Principal Component Analysis (PCA), Multivariate Linear and Logistic Regression and Person Correlation was used for complex multivariate data analysis and data stratification and among the outcome models. The p-value <0.050 at 95% confidence interval (CI) was reported as statistically significant.

## 3. Results

### 3.1. CSF leukocyte infiltration negatively correlates with CSF cryptococcal fungal burden and positively correlate with CSF protein and peripheral CD4^+^ and CD8^+^ T cell counts

We defined the baseline participant demographics, clinical, microbiologic, and immunological features that correlated with CSF leukocyte counts (Table 1 and Figure 1). Among participants, 51.5% (206/400) were antiretroviral therapy (ART) experienced, with median CD4^+^ T cells/µL of 16 (interquartile range [IQR]; 6-43). The CSF leukocytes were generally low (median of <5 [IQR; <5-45] cells/µL). The CSF cryptococcal fungal colony forming units (CFUs) were at a median of 52,000 (IQR; 1195 to 335,000) CFU/mL.

**Figure 1.**
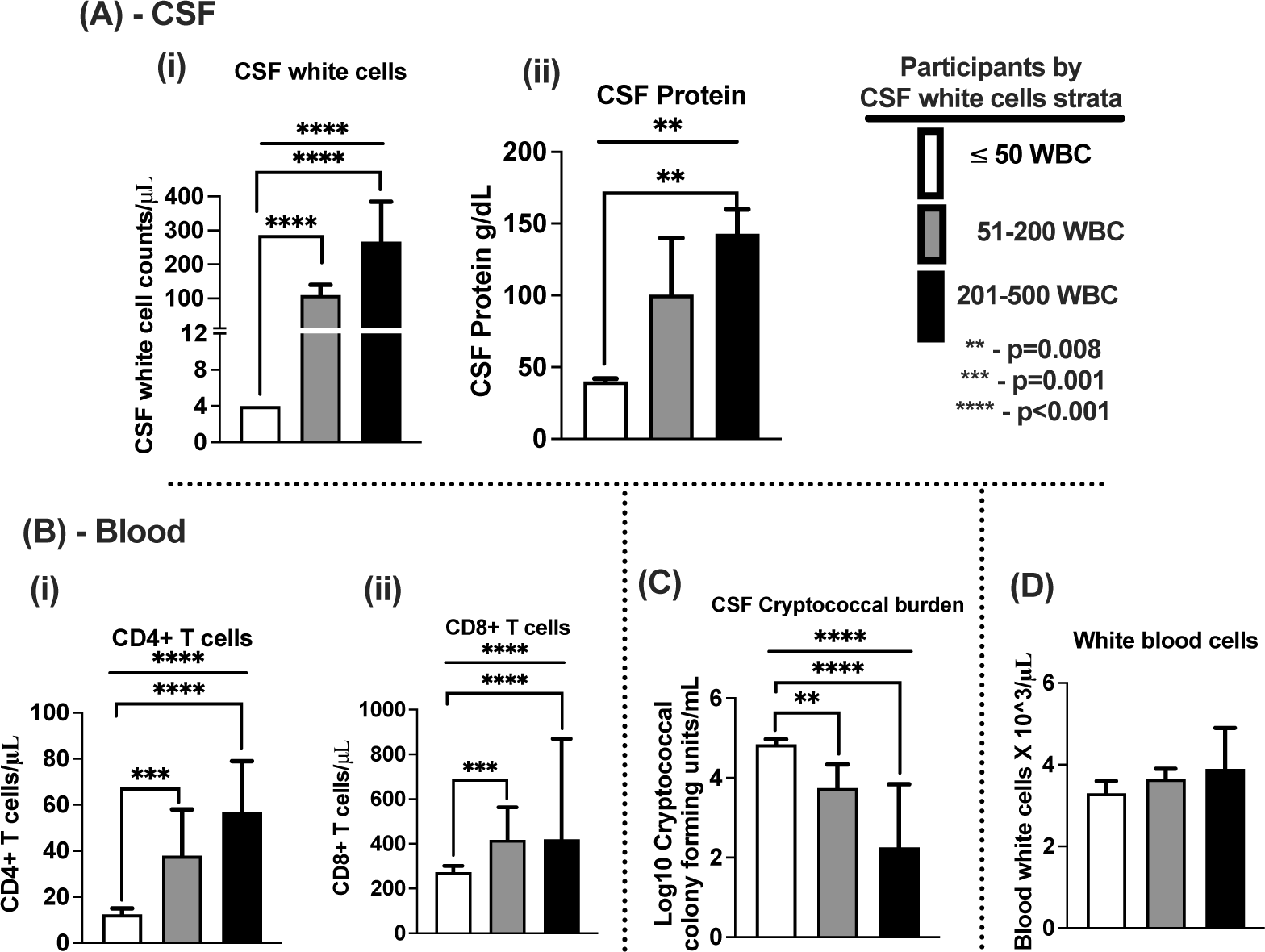
the correlation of CSF and blood clinical features. A - correlation of CSF features with 18 weeks survival by levels of CSF white cells. A (i) – CSF white cells, A (ii) – CSF proteins and A (iii) CSF cryptococcal fungal burden. B – correlation of peripheral blood features with 18 weeks survival by levels of CSF white cells. B (i) – peripheral blood white cells, B (ii) - CD4^+^ T cells, and B (iii) - CD8^+^ T cells. Error bars – median and 95% confidence intervals. * - statistically significant variables reported at p-value <0.050, at 95% confidence intervals.

**Table 1.** Participants Baseline Demographics and Clinical Features by CSF Leukocytes. Values are median (IQR). Statistic: Kruskal Wallis test (ANOVA) comparing the variables among participants across the three CSF white cell class intervals. Not statistically significant were variables with p-value ≥0.05 at a 95% confidence interval. ART* experience - antiretroviral therapy; reported among 165 participants with ≤50 cells/µL white cells, 32 participants with 51-200 white cells/µL, and 14 participants with 201-500 white cells/µL. GCS - Glasgow Coma Scale. CFU –colony forming units in CSF cryptococcal fungal growth culture. Normal adults CSF opening pressure is <10-15 mmHg that shows generally high CSF opening pressure above normal among patients with cryptococcal meningitis.

| Baseline Demographics | ≤50 white cells/μL CSF | 51-200 white cells/μL CSF | 201-500 white cells/μL CSF | P-value |
| --- | --- | --- | --- | --- |
| N | 318 | 57 | 26 |  |
| Age, years | 35 (29-41) | 32 (29-38) | 33 (28-39) | 0.227 |
| Females, n (%) | 127 (39.9) | 22 (39.6) | 11 (42.3) | 0.950 |
| Weight, kg | 51.5 (50-60) | 55 (50-60) | 50 (49.5-60) | 0.428 |
| Months on ART* | 10.0 (1.3-39.5) | 6.4 (1.2-38.6) | 2.7 (0.4-4.5) | 0.132 |
| Headache duration, days | 14 (7-30) | 14 (7.5-30) | 14 (7-23.3) | 0.924 |
| GCS <15, n (%) | 146 (67.3) | 29 (50.9) | 17 (65.4) | 0.149 |
| Hemoglobin, g/dL | 11.7 (10.1-13.3) | 11.4 (10.1-13.6) | 10.5 (9.2-12.4) | 0.127 |
| Platelets, x10<sup>3</sup>/μL | 192 (132-251) | 203 (145-254) | 225.5 (136 -329) | 0.258 |
| CSF opening pressure, mm Hg | 19.7 (14.7-22.1) | 17.7 (13.2-20.9) | 19.1 (15.3-20.1) | 0.653 |

The baseline participant demographics, clinical, and laboratory features (Table 1) and peripheral blood white blood cells (Figure 1) did not differ by CSF leukocyte stratification (Figure 1 (i)). However, both CSF protein (Figure 1A (ii)) and peripheral CD4^+^ and CD8^+^ T cells (Figure 1B (i-ii) respectively) correlated positively with CSF WBC number, whereas fungal burden was lower and correlated negatively (Figure 1C) (Pearson r, correlation data not shown). The peripheral white blood cells only had a trend to increasing with CSF white cell stratification (Figure 1 D).

### 3.2. CSF cytokines and chemokine concentration positively correlate with CSF leukocytes infiltration

Next, we determined the association of CSF cytokine and chemokine concentration in relation to CSF leukocyte infiltration. The level of Th1 cytokines (IL-2, IFN-γ and TNF-α correlated with the levels of CSF leukocytes infiltration (Figure 2A (i-iii). Similarly, levels of Th17 cytokine IL-17, the immune regulatory element, IL-10 cytokine and immune checkpoint PD-L1 correlated with CSF leukocytes infiltration (Figure 2A-C), as did the CXCR3^+^ T cell chemoattractant chemokine CXCL10/IP-10 and myeloid cells chemoattractant chemokine CCL11/Eotaxin (Figure 2C-D).

**Figure 2.**
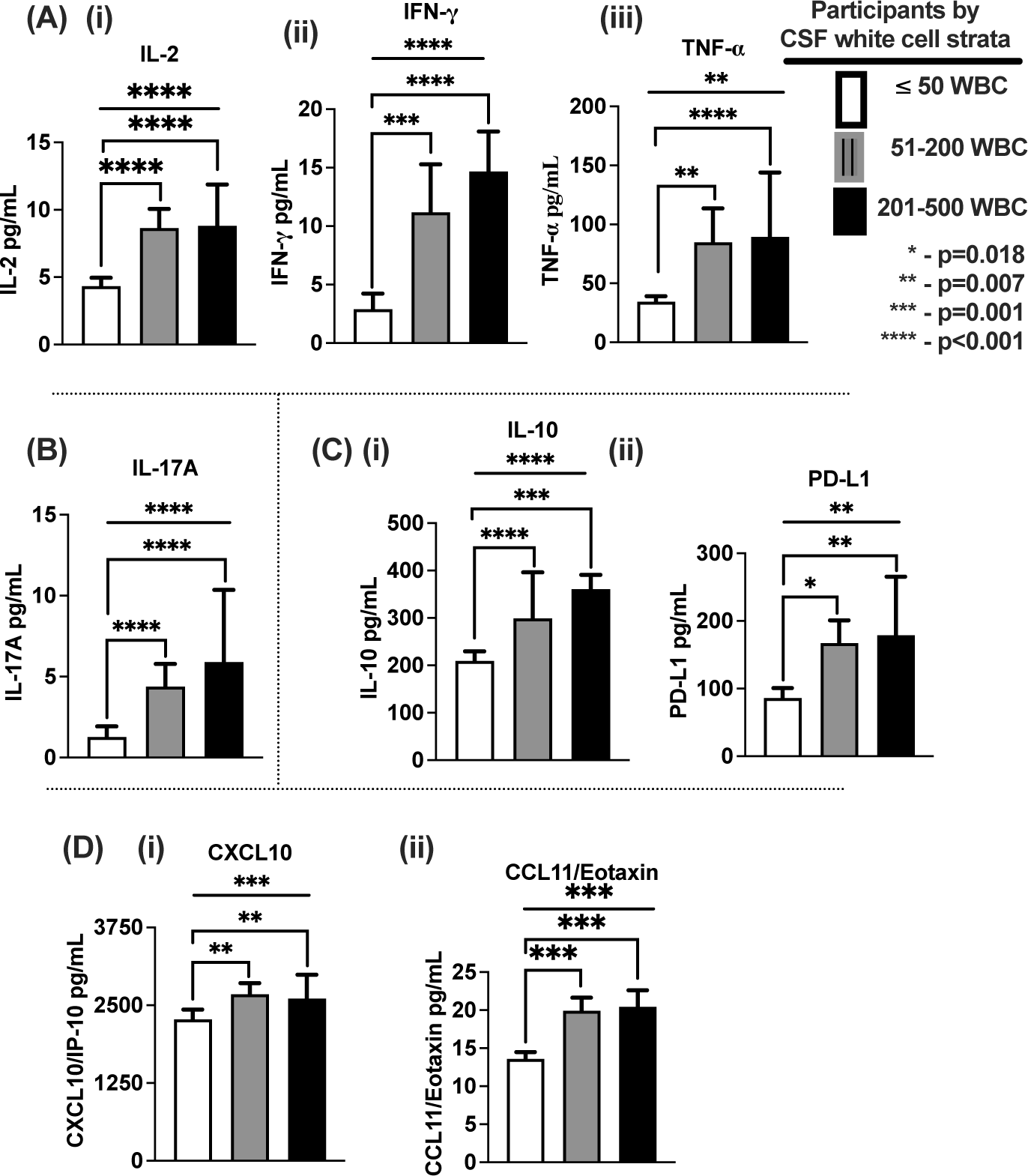
Correlation of CSF cytokines and chemokine levels with CSF leukocyte counts. A- Th1 cytokines; A (i) - Interleukin 2, A (ii) - Interferon gamma, A (iii) - Tumor necrosis factor alpha. B - Th17 cytokine, IL-17A. C – Immune regulatory elements; (i) – Interleukin 10, and programmed death 1 ligand. D Chemokines; D (i) CXCL10/IP-10 and D (ii) – CCL11/Eotaxin. The CSF white cells; (≤50 cells/μL; n=318), (51-200 cells/μL; n=57) and (201-500 cells/μL; n=26) participants. Error bars – median and 95% confidence intervals. * - statistically significant variables reported at p-value <0.050, at 95% confidence intervals.

### 3.3. Paucity of cellular and soluble immune activated response is associated with low probability of 18-week survival

The 18-week survival was 55% (203/369 participants). Survival at 18-weeks of observation was associated with CSF leukocyte infiltration (Figure 3). The probability of survival was lowest (151/292 participants; 51.7%) among individuals with CSF leukocytes ≤50 cells/μL (median <5) compared with those with 51-200 (median 110) and 201-500 (median 268) cells/uL (51% vs. 67.3% vs. 68%, respectively) (Log Rank p=0.028) (Figure 3B). This association of CSF WBC number and survival was consistent with fewer (Figure 3C) and more WBC strata (Figure 3A) (stratified adjusted Hazard Ratio, (HR= 1.634, 95% CI; 1.140 to 2.343) and p=0.008) (Figure 3C).

**Figure 3.**
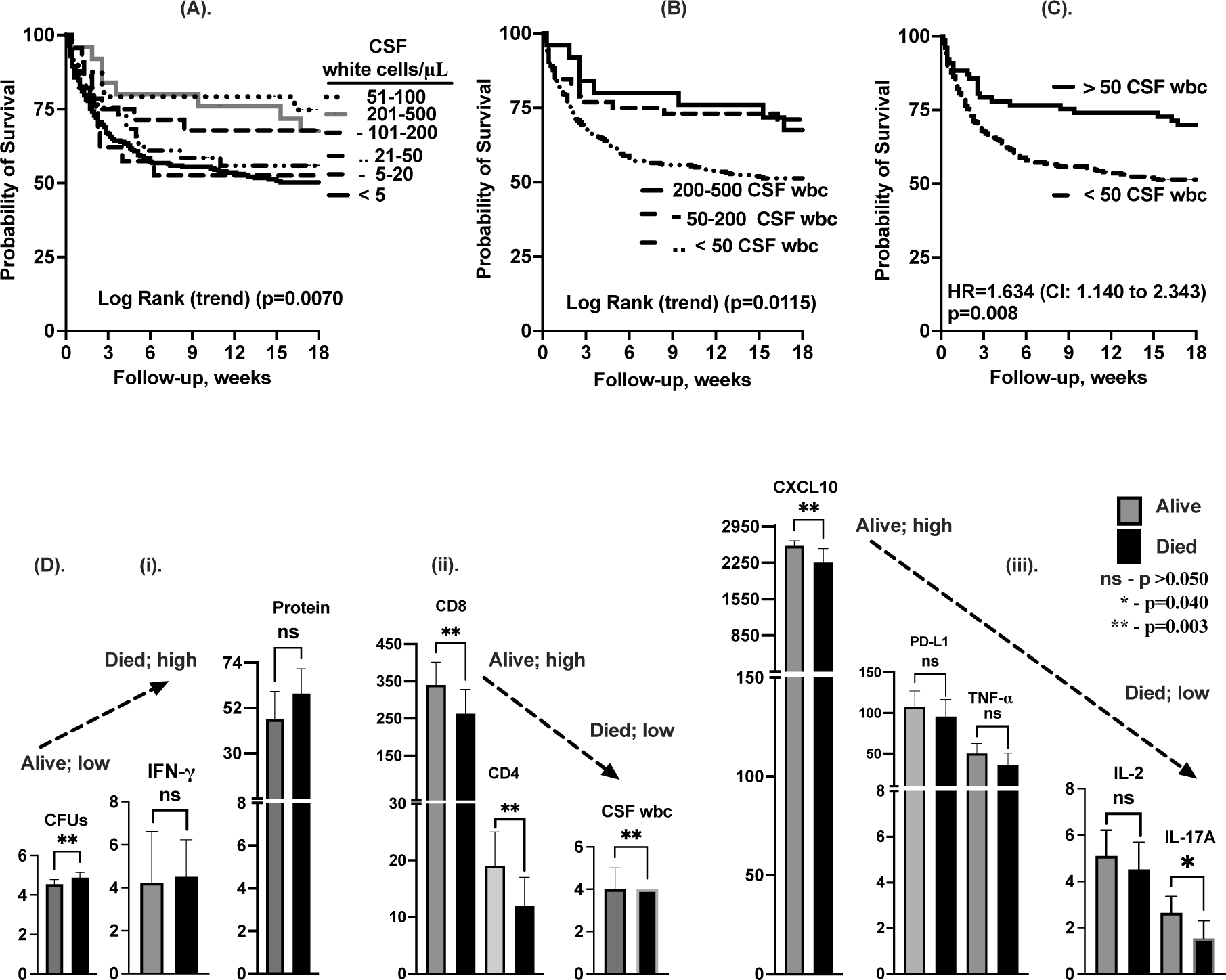
correlation of CSF white cells with 18-week survival. A –survival by CSF white cell intervals (<5; n=245), (5-20; n=31), (21-50; n=42), (51-100; n=26), (101-200; n=31), and (201-500; n=26). B - 18 weeks survival by CSF white cells; (≤50 cells; n=318), (51-200 cells/μL; n=57) and (201-500 cells/μL; n=26) participants. C – 18-week survival with CSF ≤50 cells/μL. D (i-iii) –illustrates univariate immune responses associated with survival between survivors and those who died during 18-weeks of follow-up. Statistics - Mann-Whitney unpaired t-test. * - show statically significant variables. NS-not significant. Error bars – show 95% confidence intervals. p-values, p<0.050 were statistically significant.

Survivors had significantly lower levels of CSF fungal burden/CFUs/mL, (Figure 3 D (i), high levels of CD4^+^ and CD8^+^ T cells and CSF white cells (Figure 3 D (ii)), high levels of CXCL10, and IL-17A (Figure 3 D (iii) compared to those who died. Other variables IFN-γ, CSF protein tended to be lower with survival. (Figure 3D (i)). In contrast, PD-L1, TNF-α and IL-2 tended to be high with survival (Figure 3D (iii)).

### 3.4. Principal component analysis shows clustering of CSF fungal burden, and host survival with circulating immune response

To integrate putative survival-related variables, we visualized the associated data clusters among all participants by Eigenvector projections on principal components (PC) 1 and PC2, respectively (Figure 4). The principal component analysis (PCA) showed three clusters of the cellular and soluble immune factors with fungal burden and host survival by Eigenvector projections on the PCA model (Figure 4A). A diagonal clustering was observed between CSF cryptococcal CFUs/mL and host survival (PCA cluster (i)) and with (CSF white cells, CD4^+^ and CD8^+^ T cells, white blood cells and CSF protein (PCA cluster (ii) (Figure 4A (i-ii)). Two orthogonal clusters were observed for CSF cryptococcal CFUs/mL and host survival with CSF cytokines (PCA cluster (i) and (iii); Figure 4A) and CSF cytokines with CSF and peripheral blood features (PCA cluster (ii) and (iii); Figure 4A) respectively. Cumulatively, the CSF leukocytes Eigenvector projection on PC1 had a 35.7% effect size, and the CSF cryptococcal CFUs/mL Eigenvector projection on PC2 with a 12.8% effect size, contributing the greatest variability to the PCA model (Figure 4B). The principal component with highest effect sizes are those with the most likelihood that to predict the model outcome for within component, between components and among components interactions. In this case fungal CFU and CSF WBC was demonstrated to have the greatest prediction power on survival.

**Figure 4.**
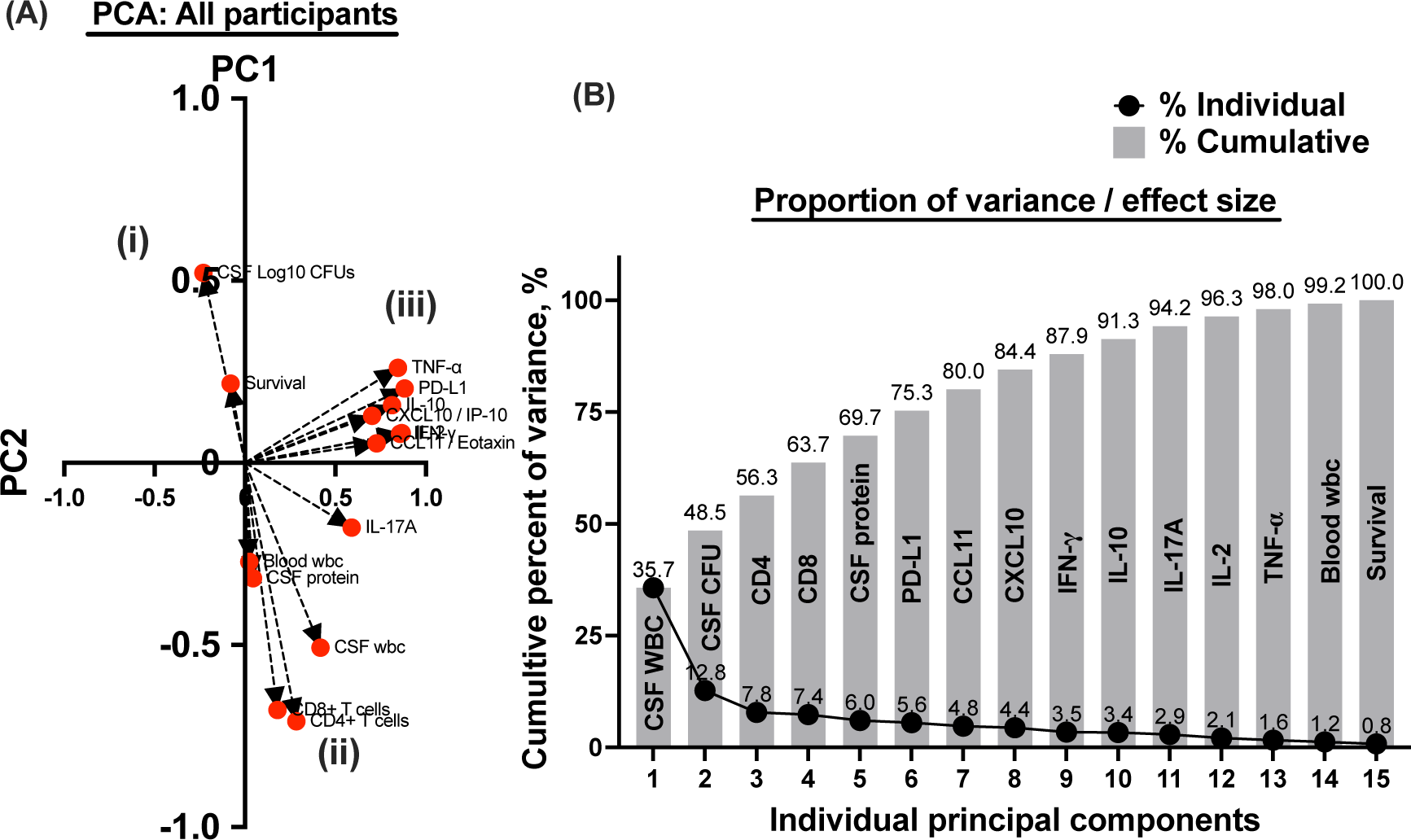
Principal Component Analyses showing Eigenvector projection (A) and cumulative effect size contributed by each covariate the PCA model (B) examined among all participants. The three planes demonstrated by the Eigenvector projects are; (i-ii) - diagonal plane between CSF fungal burden and host survival and CSF white cells, CSF protein, peripheral white cells, CD4^+^ and CD8^+^ T cells. (i-iii) orthogonal plane between CSF fungal burden and host survival and the cytokine/chemokine profile. (ii-iii) orthogonal plane between CSF fungal burden and host survival and CSF white cells, CSF protein, peripheral white cells, CD4^+^ and CD8^+^ T cells. Eigenvectors >5 shows greater power and association with the outcome variable among interacting covariates.

### 3.5. CSF leukocyte infiltration correlates with circulating CD4^+^ and CD8^+^ T cells, and CSF fungal burden and levels of IL-17A and TNF-α

Next, we determined the contribution of the cellular and soluble immune factors that independently determine CSF white cells and host survival. After adjusting for PCA components in cluster (ii) with the CSF white cell counts, CSF cryptococcal, Log_10_ CFUs/mL; (p=0.0022), CD4^+^ T cells counts; (p=0.0119), CD8^+^ T cells counts; (p=0.0015), and CSF protein, mg/dL; (p=0.0143), each independently predicted CSF leukocyte infiltration among all patients, (Table 2; Model 1). This observation was consistent even with substitution of 7.9% (32 of 401) missingness in survival (all alive or all dead) (data not shown). After adjusting for PCA components in cluster (iii) with the levels of CSF leukocyte infiltration, the levels of IL-17A; (p<0.0001) a Th17 cytokine, and TNF-α; (p=0.0261) a Th1 cytokine, independently predicted the levels of CSF leukocyte infiltration among all patients, (Table 2; Model 2).

**Table 2.** Multivariate Linear Regression of Independent Predictors of CSF Leukocytes Infiltration. Statistic: Multivariate Linear Regression (least squares) models among all participants. Cytokines; (pg/mL). The values (Risk Ratios) - shown model transformed and normalized imputed values (not actual measurements) of variables. P values, p<0.0500 are statistically significant. Other variables including peripheral white blood cells, PD-L1, CXCL10, CCL11, IFN-γ, IL-2 and IL-10 did not independently predict the levels of CSF white cells with cryptococcal meningitis.

| Independent adjusted variables | Estimated Risk Ratios (95% CI; asymptotic) | Dependent | P value |
| --- | --- | --- | --- |
| Model 1: (PC1; (ii)) CSF white cell dependent and matched independent colinear predictor variables. Adjusted for PCA cluster (ii) components; (CSF cryptococcal, log10 CFUs/mL, CD4<sup>+</sup> T cells /μL, CD8<sup>+</sup> T cells /μL, CSF protein, mg/dL, and blood white cells /μL). |  |  |  |
| CSF cryptococcal, log10 CFUs/mL | -8.11 (-13.28 to -2.94) | CSF white cells | 0.002 |
| CD4<sup>+</sup> T cells /μL | 0.28 (0.06 to 0.50) | CSF white cells | 0.012 |
| CD8<sup>+</sup> T cells /μL | 0.05 (0.02 to 0.078) | CSF white cells | 0.002 |
| CSF protein, mg/dL | 0.09 (0.02 to 0.16) | CSF white cells | 0.014 |
| Model 2: PC1; (iii) CSF white cell dependent and matched independent colinear predictor variables. Adjusted for PCA cluster (iii) components; (B7-H1 / PD-L1, CCL11 / Eotaxin, CXCL10 / IP-10, IFN-γ, IL-10, IL-17A, IL-2 and TNF-α) |  |  |  |
| IL-17A | 3.04 (1.97 to 4.12) | CSF white cells | <0.0001 |
| TNF-α | -0.16 (-0.29 to -0.02) | CSF white cells | 0.026 |

The CSF white cell infiltration during cryptococcal meningitis (cryptococcal CFU/mL) is associated with the number of peripheral circulating CD4^+^ and CD8^+^ T cells and concentration of CSF protein. These immune modulators together predict the level of IL-17A and TNF-α cytokine concentration in CSF with cryptococcal meningitis.

### 3.6. PD-L1, CXCL10 and IL-2 independently predict survival

After adjusting for all components in the PCA outcome model, elevation in levels of B7-H1 / PD-L1 in both linear and logistic regression analysis consistently predicted higher probability of survival (Table 3; Model 1). The higher levels of CXCL10 / IP-10 (model 1-2) consistently and independently predicted higher probability of survival among all patients. (Table 3; Model 1). This observation was consistent even after adjusting for 7.9% missingness in survival (data not shown). The elevation in the levels of IL-2 in both linear and logistic regression analysis (Model 1-2) consistently and independently predicted higher probability of survival among all patient (Table 3; Model 1-2) respectively. Each of the soluble immune factors IFN-γ, IL-2, TNF-α, and IL-17A cytokines and CXCL10 / IP-10 chemokine were independently associated with levels of PD-L1, (in all factors multivariate linear regression analysis p<0.020 and data not shown).

**Table 3.** Multivariate Linear and Logistic Regression of Independent Predictors of Survival. Statistic: Multivariate Linear Regression (Least Squares) and Multivariate Logistic Regression Survival outcome dependent predictor variables among all participants. Interpretation: the values (Risk Ratios and Odds Ratios) - shown model transformed and normalized imputed values (not actual measurements) of variables. In Linear regression, the p-values, p<0.0500 were statistically significant. CI – confidence interval. The confidence interval values not crossing through zero on either direction (positive or negative) are statistically significant. The PCA cluster (ii) adjusted components including (CSF white cells, CSF cryptococcal log10 CFUs, CD4^+^ T cells, CD8^+^ T cells, CSF protein, and blood white cells) and other variables did not predict or match survival in either analyses (model analysis data not shown).

| Independent adjusted variables | Estimated Risk Ratios (95% CI) | Linear regression, P value | Logistic regression; Odds Ratios (95% CI) |
| --- | --- | --- | --- |
| Model 1: All Principal Components survival dependent and matched independent predictor of survival. Adjusted for CSF white cells /μL, CSF cryptococcal log10 CFUs /mL, CD4 T cells /μL, CD8<sup>+</sup> T cells /μL, CSF protein, mg/dL, B7-H1 / PD-L1, CCL11 / Eotaxin, CXCL10 / IP-10, IFN-γ, IL-10, IL-17A, IL-2, TNF-α and blood white cells /μL. | | | |
| B7-H1 / PD-L1 | 0.0010 (4.1e-005 to 0.0021) | 0.0414 | 1.0060 (1.0010 to 1.0110) |
| CXCL10 / IP-10 | -0.0001 (-0.0001 to -5.4e-006) | 0.0320 | 0.9997 (0.9995 to 1.0000) |
| IL-2 | 0.0280 (0.0054 to 0.0507) | 0.0155 | 1.1430 (1.0330 to 1.2720) |
| Model 2: PC2: PCA cluster (iii) survival dependent and matched independent predictor variables. Adjusted for (B7-H1 / PD-L1, CCL11 / Eotaxin, CXCL10 / IP-10, IFN-γ, IL-10, IL-17A, IL-2 and TNF-α) | | | |
| CXCL10 / IP-10 | -0.0001 (-0.0001 to -2.1e-005) | 0.0045 | 0.9997 (0.9995 to 0.9999) |
| IL-2 | 0.0207 (0.0014 to 0.0399) | 0.0353 | 1.0920 (1.0080 to 1.1870) |

## 4. Discussion

Survival with HIV-associated cryptococcal meningitis is associated with lower CSF fungal burden, greater CSF leukocyte infiltration and higher levels of neuroimmune factors in CSF. We found that individuals with CSF leukocytes <50 cells/μL had the poorest clinical and immunological profile associated with the highest CSF fungal burden, lowest number of peripheral CD4^+^ and CD8^+^ T cells, CSF cytokines and the lowest probability of 18-week survival than individuals compared with those with >50 CSF leukocytes/µL. Among cytokines in CSF, individuals with CSF leukocytes <50 cells/μL had the lowest levels of the Th1 cellular growth activating cytokine IL-2, cellular activating cytokines IFN-γ, cell death activating cytokine TNF-α, and CXCR3^+^ T cell chemoattractant chemokine CXCL10 and Th17 T cell-activating cytokine IL-17A. In multivariate analysis, infiltrating CSF leukocyte numbers were negatively associated with CSF cryptococcal fungal burden, but positively associated with CSF protein concentration and peripheral circulating CD4^+^ and CD8^+^ T cells. Moreover, elevated concentrations of CXCL10, IL-2 and PD-L1 independently predicted a higher probability of 18-week survival.

In cryptococcal meningitis, nearly 80% of the baseline CD45^+^ WBC in CSF are T cells, among which 70% are CD8^+^ and a minority are CD4^+^ [8]. Other mononuclear cells central to the CSF exudate included NK cells, monocytes [8] and B cells [7]. Critical to the outcome of cryptococcal infection is the balance of an activated cellular immune response with a poly-functional soluble immune profile that function in synergy [29–31]. The primary cells infected are tissue resident macrophages that provide an intracellular niche for cryptococcal replication [29–31]. To prevent intracellular cryptococcal replication requires an activated T cell response. Cytotoxic CD8^+^ T cells and NK T cells are modulated by Th1 CD4^+^ T cell stimulation [32]. It is notable that evasion of multiple components of innate and adaptive immune system collectively predispose hosts to virulent and lethal cryptococcal disease [29–31]. In this regard, the immune damage response framework suggests that antifungal therapy alone without immune modulatory therapy is not sufficient to cure the infection and modulate host immune-mediated injury and death.–Thus, multivariate linear and logistic regression models that integrate cytokine, chemokine, and cellular responses, as described herein, reveal the potential role of poly-functional immune modulated responses to influence fungal burden and host survival.

The inverse association of the CSF fungal burden with the levels of cellular and soluble immune response and host survival implicates the selective evasion of Th1- and Th17-mediate control of intracellular *Cryptococcus* replication. We showed that upregulation of IL-17A is associated with lower CSF cryptococcal fungal burden and improved survival. In the mouse lung model, elevated IL-17 modulates cryptococcal yeast differentiation to limit spread across the blood-brain barrier [33–35]. The adaptive immune clearance of *Cryptococcus* requires precise activation and recruitment of cytokine- and chemokines-producing T cells that serve as the cornerstones of protection[6]. The evasion of cryptococcal fungal immune control mechanisms with down-regulated Th1 (IFN-γ, TNF-α, IL-2) and Th17 T cell cytokines (IL-17A) and CXCR3^+^ T cell chemoattractant chemokine (CXCL10) responses, allows more vigorous replication of *Cryptococcus* and overwhelming infection. Here we report that fatal instances of cryptococcal meningitis are associated with a paucity of CSF CXCR3^+^ T cell activating chemokine CXCL10, growth activating cytokine IL-2 and immune checkpoint regulatory element PD-L1. The down regulated expression of these neuroimmune modulatory molecules in cryptococcal disease selectively likely impairs recruitment and maturation of adaptive T cell function in response to the CNS intracellular fungal replication.

In this context, *Antonia et al., (2019)* showed that low concentrations of CXCL10 in circulation were associated with glycoprotein proteolytic cleavage of the CXCL10 chemokine terminal domain by glycoprotein[33]. The effect of the extracellular cleavage of CXCL10 ligand also affected other members of the CXCR3^+^ receptor family of activating chemokines. These members include CXCL11 and CXCL9 that impair T cell recruitment in response to intracellular evading class of pathogens [33]. Consistent with our results, patients with low levels of CSF CXCL10 and a low probability of survival demonstrate impaired fungal clearance [34] with a high fungal burden [12]. These observations are consistent with a T cell evasion hypothesis in which impaired immune control of intracellular cryptococcal replication is associated with paucity of T cell activated response [31]. Th1 T cell produce IFN-γ, TNF-α IL-2 activation cytokines and CXCR3^+^ T cells CXCL10 chemoattractant enable infiltration of CSF by T, NK, NK-T and myeloid cell lineages [35]. The decrement in CXCR3^+^ receptor family chemoattractant chemokines impairs CSF T cell recruitment and cytokine activation at the site of infection, which allows intracellular replication of *Cryptococcus* resulting in high fungal burden and impaired host recovery.

Among Th-1 cytokines, induction of IFN-γ facilitates activation and recruitment of effector cellular responses needed to control infection [43]. IFN-γ induces the chemokine CXCL10/IP-10, a chemoattractant ligand which recruits activated lymphocytes in response to CNS infection [37,38]. The disruption of IFN-γ-encoding genes increased the susceptibility to infection and rapid progression to death [36,39]. Whether mutations in IFN-γ encoding genes observed in other infections contribute to the high in hospital mortality rates with Cryptococcal infection has not been investigated [12,13]. Barber et al., demonstrated the importance of IFN-γ responses in modulating immune activation among intracellular *M. tuberculosis*-infected macrophages [40]. The absence of an IFN-γ activating response limited production of reactive oxygen species needed to fully activate intracellular/phagosome killing in infected macrophages with increased mycobacterial burden [40].

Clinically, the low peripheral CD4^+^ T cell count despite ART treatment at CM diagnosis potentially implicates the role of ART failure or failed immune reconstitution with unmasking and/or progression of cryptococcal infection. The use of high dose of antifungals early can constitute effective pre-emptive therapy to improve outcomes before patients get severely ill with high CSF cryptococcal fungal burden. However, early diagnosis or staging of cryptococcal infection is a challenge where patients present late with overt disease 1-2 weeks after the onset of symptoms [13,41].

## 5. Conclusion

Multiple cellular and soluble immune elements appear to mediate adaptive-innate immune crosstalk to control intracellular cryptococcal replication and modulate host recovery. A paucity of CSF white cells is associated with higher fungal burden, lower levels of neuroimmune cytokines, chemokines and checkpoint inhibitors and a lower frequency of survival at 18-weeks. The decrement in these factors likely conspire to limit T cell-mediated control of intracellular cryptococcal replication in macrophages with resultant serious infection and death. The levels of evoked Th1 and Th17A cytokines and CXCR3^+^ activating chemokine CXCL10 and PD-L1 immune checkpoint responses are potentially modifiable to enhance clinical outcomes. These variables also serve as diagnostic, prognostic, and survival indicators required to advance immune based therapy to control *Cryptococcus* and improve survival. Given that antifungal therapy alone is not sufficient to cure cryptococcal meningitis. Immune modulatory treatment may hold promise going forward.

## Data Availability

All data produced in the present study are available upon reasonable request to the authors.

## 8. Author contribution

SO - conceptualized the study, framed the hypothesis, designed statistical models, performed statistical analyses, compiled the results, drafted the first version of the manuscript, edited the manuscript, managed the publication review process, and sourced and contributed research funding. DBM - conceptualized the study, framed the hypothesis, supported experiment design, guided data analysis, guided manuscript writing, contributed to the first draft version of the manuscript, edited the manuscript, sourced, and contributed research funding. ENJ - conceptualized the study, framed the hypothesis, guided data analysis, guided manuscript writing, edited the manuscript, sourced, and contributed research funding. YCM - conceptualized the study, framed the hypothesis, edited the manuscript, and sourced and contributed research funding. DRB - designed parent cohort studies, guided data analysis, edited the manuscript, sourced, and contributed research funding. JR - conceptualized the study, framed the hypothesis, edited the manuscript contributed to the research funding. OOJ - conceptualized the study, framed the hypothesis, and edited the manuscript. LT, CPS EK, AKM, ECO, managed the patients, contributed data, and reviewed the manuscript. AA managed patients’ diagnostics, contributed data, and reviewed the manuscript.

## Acknowledgments

We thank study participants for their involvement in the parent study. We thank the ASTRO trial team for the clinical management of patients and data collection including Jane Francis Ndyetukira, Cynthia Ahimbisibwe, Florence Kugonza, Alisat Sadiq, Catherine Nanteza, Kiiza Tadeo Kandole, Darlisha Williams, Edward Mpoza, Apio Alison, Radha Rajasingham, Mahsa Abassi, Enock Kagimu, Morris K. Rutakingirwa, Fiona Cresswell, and John Kasibante. We that the doctorial committee mentorship support from Makerere University, especially from Bernard S. Bagaya, Freddie Bwanga, and Henry Kajumbula. We thank institutional support from IDI from Bosco Kafufu, Andrew Kambugu. We thank the Ph.D. program mentorship support received from; the IDI Research Capacity Building Unit especially from Barbara D. Castelnuovo, Aidah Nanvuma, Stephen Okoboi, and the Statistics unit especially Agnes Kiragga. John Hopkins University, School of Medicine and Bloomberg School of Public Health, Department of Molecular Microbiology, and Immunology, especially from Robert R. Bollinger and Arturo Casadevall. From the University of Colorado, Denver, Anschutz Medical Campus Aurora, Division of Infectious Disease, Department of Medicine and Veteran Affairs Research Services especially from Brent Palmer, Tina Powell, and Jeremy Rakhola. We thank the statistical data analysis mentorship received from the University of Minnesota, especially from Ananta S. Bangdiwala and Kathy Huppler Hullsiek.

